# Preliminary Analysis of Safety and Immunogenicity of a SARS-CoV-2 Variant Vaccine Booster

**DOI:** 10.1101/2021.05.05.21256716

**Authors:** Kai Wu, Angela Choi, Matthew Koch, LingZhi Ma, Anna Hill, Naveen Nunna, Wenmei Huang, Judy Oestreicher, Tonya Colpitts, Hamilton Bennett, Holly Legault, Yamuna Paila, Biliana Nestorova, Baoyu Ding, Rolando Pajon, Jacqueline M Miller, Brett Leav, Andrea Carfi, Roderick McPhee, Darin K Edwards

## Abstract

Severe acute respiratory syndrome coronavirus 2 (SARS-CoV-2) is the causative agent of a global pandemic of coronavirus disease 2019 (COVID-19) that has led to more than 3 million deaths worldwide. Safe and effective vaccines are now available, including the mRNA-1273 prototype vaccine, which encodes for the Wuhan SARS-CoV-2 spike (S) protein stabilized in the prefusion conformation by 2 proline substitutions. This vaccine showed 94% efficacy in prevention of symptomatic COVID-19 disease in a phase 3 clinical study. Recently, SARS-CoV-2 variants have emerged, some of which have shown decreased susceptibility to neutralization by vaccine-induced antibody, most notably the B.1.351 variant, although the overall impact on vaccine efficacy remains to be determined. In addition, recent evidence of waning antibody levels after infection or vaccination point to the need for periodic boosting of immunity. Here we present the preliminary evaluation of a clinical study on the use of the prototype mRNA-1273 or modified COVID-19 mRNA vaccines, designed to target emerging SARS-CoV-2 variants as booster vaccines in participants previously vaccinated approximately 6 months earlier with two doses of the prototype vaccine, mRNA-1273. The modified vaccines include a monovalent mRNA-1273.351 encoding for the S protein found in the B.1.351 variant and multivalent mRNA-1273.211 comprising a 1:1 mix of mRNA-1273 and mRNA-1273.351. As single 50 µg booster vaccinations, both mRNA-1273 and mRNA-1273.351 had acceptable safety profiles and were immunogenic. Antibody neutralization titers against B.1.351 and P.1 variants measured by SARS-CoV-2 pseudovirus neutralization (PsVN) assays before the booster vaccinations, approximately 6 to 8 months after the primary series, were low or below the assay limit of quantification, although geometric mean titers versus the wild-type strain remained above levels likely to be protective. Two weeks after the booster vaccinations, titers against the wild-type original strain, B.1.351, and P.1 variants increased to levels similar to or higher than peak titers after the primary series vaccinations. Although both mRNA-1273 and mRNA-1273.351 boosted neutralization of the wild-type original strain, and B.1.351 and P.1 variants, mRNA-1273.351 appeared to be more effective at increasing neutralization of the B.1.351 virus versus a boost with mRNA-1273. The vaccine trial is ongoing and boosting of clinical trial participants with the multivalent mRNA-1273.211 is currently being evaluated.

## Introduction

Emergence of the severe acute respiratory syndrome coronavirus-2 (SARS-CoV-2) in December 2019 resulted in a pandemic of coronavirus disease 2019 (COVID-19) that has caused millions of deaths globally (1, 2). Vaccines for SARS-CoV-2 targeting the spike (S) protein (mRNA-1273; BNT162b2; AD26.COV2.S; AZD1222) have been developed and administered to over a billion people worldwide (3). Although these vaccines are highly effective in reducing symptoms of COVID-19 and severe disease, several viral variants with changes in the S protein and elsewhere in the viral genome have arisen, some of which have been identified as variants of concern (VOC) due to evidence of increased transmissibility and disease severity, and decreased neutralization by antibodies generated during previous infection or vaccination (B.1.1.7, B.1.351, P.1, B.1.427, and B.1.429) in the United States (US) (4). The B.1.1.7 variant which first emerged in the United Kingdom in September 2020 has become the most prevalent circulating variant in the US and in most other countries (5, 6). The B.1.351 variant is most prevalent in South Africa (∼90%) and the P.1 variant is most prevalent in Brazil (>90%) as of late March, 2021 (5). However, the overall impact of variants on vaccine efficacy is being evaluated (7).

mRNA-1273, a novel lipid nanoparticle-encapsulated messenger RNA encoding a prefusion stabilized S protein of the Wuhan-Hu-1 isolate, demonstrated anti-SARS-CoV-2 immune responses in phase 1 (NCT04283461) and 2 (NCT04405076) trials in adults, and 94% efficacy and an acceptable safety profile against symptomatic COVID-19 disease in the phase 3 Coronavirus Efficacy (COVE) (NCT04470427) trial in over 30,000 participants (8-11). The U.S.

Food and Drug Administration granted Emergency Use Authorization (EUA) in December, 2020 for the mRNA-1273 vaccine (12), the European Medicines Agency granted a conditional marketing authorization for the mRNA-1273 vaccine on January 2021 (13), and the World Health Organization has issued Emergency Use Listing for the mRNA-1273 vaccine to prevent COVID-19 in individuals 18 years of age and older on April 2021 (14).

To monitor for VOC, neutralizing capacity of clinical sera is routinely tested against variants. Antibodies from mRNA-1273 vaccinees neutralized the B.1.1.7 variant to a similar extent as the original Wuhan-Hu-1 isolate and the D614G variant; whereas neutralizing antibody titers against the P.1 variant were reduced by 3.5-fold and titers against the B.1.351 variant were reduced by 6.4-fold (15). More recently, data from the ongoing phase 1 trial showed that vaccine antibody responses persisted up to 6 months following the second dose (16). Although a neutralizing antibody titer threshold predictive of protection from SARS-CoV-2 infection is not currently known, the reduction in *in vitro* neutralizing antibody titers against some variants relative to the prototype strain, raises the possibility of breakthrough infections and waning efficacy for current SARS-CoV-2 vaccines. One approach to address variants is to develop new vaccines tailored to the variants. In mice, the initial evaluation of mRNA-1273 vaccines (mRNA-1273.351 and mRNA-1273.211) designed to target emerging SARS-COV-2 variants increased neutralizing titers against variants and showed that a booster dose significantly increased neutralizing titers against the wild-type isolate (D614G) and the variants (17). Both mRNA-1273.351 and mRNA-1273.211 vaccines are currently being evaluated in additional pre-clinical challenge models and in clinical studies. Here we report upon the preliminary safety and immunogenicity of single booster doses of mRNA-1273 and mRNA-1273.351 in a phase 2 clinical trial.

## Methods

### Study Design

The phase 2 mRNA-1273-P201 study (NCT04405076) enrolled adult participants ≥ 18 years of age at 8 sites in the U.S. Preliminary safety and immunogenicity results following two doses of 50 or 100 µg of mRNA-1273 have been previously reported (8, 9). Given that the primary efficacy endpoint for mRNA-1273 against COVID-19 was met in the phase 3 COVE trial and that EUA for the vaccine was granted, both the phase 2 and 3 trial protocols were amended to include open-label interventional phases offering participants in the placebo arm an option to receive mRNA-1273 vaccine at the end of the blinded phases. The phase 2 study offered participants previously primed with two doses of either 50 or 100 ug of mRNA-1273 an option to provide a nasopharyngeal swab for RT-PCR for SARS-CoV-2 and a blood sample for serology and immunogenicity, and to receive a single booster of a 50 μg dose of mRNA-1273. A rollover part was added to the phase 2 study, in which participants from the phase 3 COVE trial who completed a two vaccinations series of 100 ug doses of mRNA-1273 and provided a blood sample for serology and immunogenicity at day 1 received a single booster of either 20 µg or 50 µg doses of mRNA-1273.351 or 50 µg of the multivalent mRNA-1273.211 (Figure 1). In this manuscript, we present the results after a booster dose of 50 µg of mRNA-1273 (Part B) or 50 µg of mRNA-1273.351 (Part C, Cohort 1).

**Figure 1:**
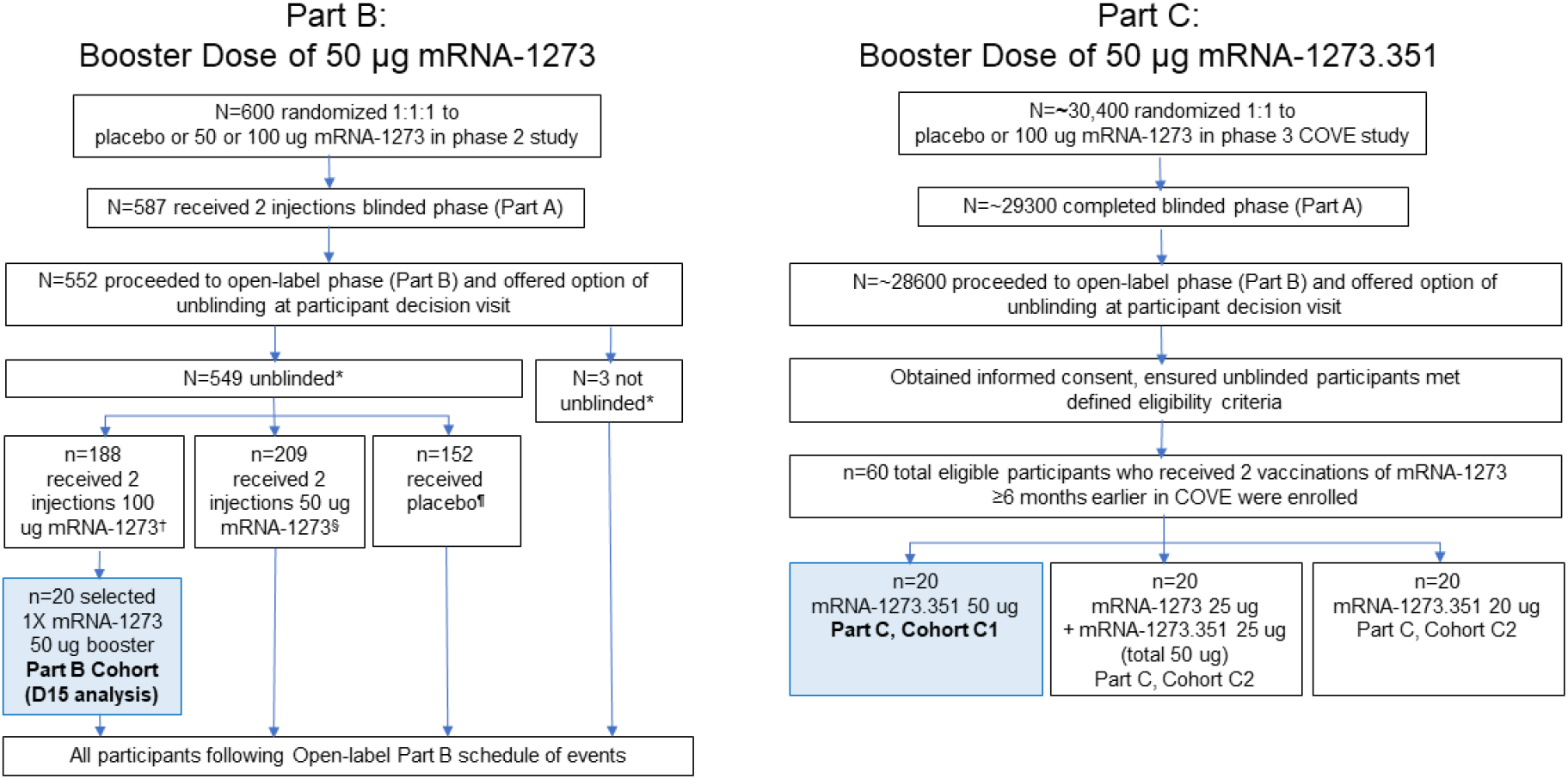
Open-label Boosting with mRNA-1273 and mRNA-1273.351 (Parts B and C) Flow of trial for Parts B and C of the amended phase 2 trial of mRNA-1273. *Unblinded or not unblinded to assigned treatment in Part A blinded phase. †Received 2 injections of 100 ug mRNA-1273 and §50 ug mRNA-1273 during Part A. ¶Received placebo in Part A. participants who had received two injections of 100 ug mRNA-1273 completed the blinded phase (Part A) of the study through the participant decision visit or were unblinded or discontinued the study. In Part B, 20 participants who had received two injections of 100 ug mRNA-1273 and completed the blinded phase (Part A) and went on to receive a single open-label booster dose of 50 ug mRNA-1273 were selected for this preliminary analysis, with selection based on completion of their D15 visit assessments and immunogenicity sample availability. In Part C, enrollment and administration of booster doses in each study arm occurred in a sequential manner. Blue boxes designate cohort data included in this report.

In Part B, all participants in the phase 2 mRNA-1273 study who previously received two doses of 50 ug or 100 µg of mRNA-1273 and provided a written consent to participate in this Part B, were administered a booster dose of 50 µg of mRNA-1273 vaccine at 177 to 226 days (5.9 to 7.5 months) after receiving the second dose of mRNA-1273. Upon enrollment into Part C of this amended phase 2 study, eligible participants from the COVE study received a single intramuscular injection of mRNA-1273.351 (20 μg or 50 μg) or mRNA-1273.211 (50 μg total) at 168 to 198 days (5.6 to 6.6 months) after receiving the second vaccination of mRNA-1273 (Figure 1). The enrollment and vaccination in each study arm cohort of Part C was sequential. The first 20 participants who gave consent to participate were enrolled and dosed on open-label day 1 (OL-D1) with the 50 μg dose of mRNA-1273.351. Upon completion of the first cohort with 50 μg of mRNA-1273.351, 20 participants were enrolled in the second cohort and received the 50 μg dose of mRNA-1273.211. Following completion of the second cohort, 20 participants were enrolled and dosed with the 20 μg dose of mRNA-1273.351.

### mRNA-1273 and mRNA-1273.351 Vaccines

The mRNA-1273.351 vaccine, like mRNA-1273, encodes the prefusion stabilized S protein of SARS-CoV-2 with the key amino acid changes present in the B.1.351 strain of the virus. The amino acid changes in the S protein encoded by mRNA-1273.351 relative to Wuhan-Hu-1 were L18F, D80A, D215G, Δ242-244, R246I, K417N, E484K, N501Y, D614G, and A701V. mRNA-1273.211 is a 1:1 mix of 25 µg of mRNA-1273 and 25 µg of mRNA-1273.351, for a total dose of 50 µg of mRNA. All vaccines were formulated in lipid nanoparticles as previously described (11).

### Inclusion and Exclusion Criteria

Eligible participants were adults, 18 years of age, considered by the investigator to be healthy at screening and day 1 of the open-label phase. Part B participants must have been previously enrolled in the mRNA-1273 P201 study. For Part C, participants must have been previously enrolled in the mRNA-1273 COVE study and received two doses of mRNA-1273 in Part A of that study (i.e., already unblinded and aware of their actual treatment), with their second dose at least 6 months prior to enrollment in Part C and must have been currently enrolled and compliant in that study (i.e., not have withdrawn or discontinued early).

Additionally, eligible participants in Part C included healthy adults or adults with pre-existing medical conditions who were in a stable condition (disease not requiring significant change in therapy or hospitalization for worsening disease during the 3 months before enrollment). Participants in the phase 3 mRNA-1273 COVE study must have provided SARS-CoV-2 serology samples at the Participant Decision Visit (OL-D1) and days 29, and 57. Exclusion criteria for Parts B and C included acute illness or febrile (≥ 38.0°C/100.4°F) 24 hours prior to or at the screening visit (day 0), current treatment with investigational agents for prophylaxis against COVID-19, receipt of systemic immunoglobulins or blood products within 3 months prior to screening, positivity for SARS-CoV-2 by RT-PCR at baseline or at any time during the mRNA-1273 COVE study regardless of the presence or absence of symptoms consistent with COVID-19, or experienced any serious adverse event in the mRNA-1273 COVE study. Pregnant or breastfeeding females, and sexually active males and females unwilling to use adequate contraception for at least 3 months after the second study vaccination were also excluded. See additional eligibility criteria (clinicaltrials.gov NCT04405076).

### Assessment of Safety

In order to assess safety, participants completed an electronic diary to record solicited systemic and local adverse reactions, daily oral body temperatures, injection site erythema and swelling/induration. Trained site personnel made telephone calls to the participants to assess safety every 4 weeks.

### Assessment of Immunogenicity

For assessment of immunogenicity, serum samples taken from participants on days 1, 8, 15, 29, 57 and 181 were analyzed for serum neutralizing antibody against SARS-CoV-2 as measured with recombinant VSV-based pseudoviruses using the D614G, B.1.351 and P.1 variant sequences of SARS-CoV-2 S protein. For this early report, only data for day 1 and day 15 samples are included. The amino acid changes in the B.1.351 S protein relative to Wuhan-Hu-1 were L18F, D80A, D215G, Δ242-244, R246I, K417N, E484K, N501Y, D614G, and A701V. The 12 amino acid changes in the P.1 S protein relative to Wuhan-Hu-1 were L18F, T20N, P26S, D138Y, R190S, K417T, E484K, N501Y, D614G, H655Y, T1027I and V1176F. Of note the key amino acid changes K417T/N, E484K, and N501Y are shared between the B.1.351 and P.1 variants.

### Neutralizing Antibody Assay

To perform the recombinant VSV-based pseudovirus neutralization assay, codon-optimized full-length spike protein of the D614G, B.1.351 and P.1 variant sequences were cloned into pCAGGS vector. To make SARS-CoV-2 full-length spike pseudotyped recombinant VSV-ΔG-firefly luciferase virus, BHK-21/WI-2 cells (Kerafast, EH1011) were transfected with the spike expression plasmid and subsequently infected with VSVΔG-firefly-luciferase as previously described (18). For the neutralization assay, serially diluted serum samples were mixed with pseudovirus and incubated at 37°C for 45 minutes. The virus-serum mix was subsequently used to infect A549-hACE2-TMPRSS2 cells for 18 hours at 37 °C before adding ONE-Glo reagent (Promega E6120) for measurement of luciferase signal (relative luminescence unit; RLU). The percentage of neutralization was calculated based on RLU of the virus only control, and subsequently analyzed using four-parameter logistic curve (Prism 8).

### Statistical Analysis

Geometric mean titer (GMT) and geometric mean fold rise (GMFR) were calculated based on the log-transformed titers. Calculation of 95% confidence intervals (CI) was based on the t-distribution of the log-transformed titers or the difference in the log-transformed titers for GMT and GMFR, respectively, then back transformed to the original scale.

Wilcoxon matched-pairs signed rank test was used for the participant’s sample comparison, such as pre-vs post-booster doses within the same type of assay, and cross assay comparison at the same timepoint.

## Results

### Disposition of Participants

As described previously, between May 22, 2020 and July 8, 2020, 600 participants were randomized to placebo or 50 or 100 ug of mRNA-1273 administered as two injections in the phase 2 mRNA-1273 trial (9). By April 30, 2021, 188 participants who had received two injections of 100 ug mRNA-1273 completed the blinded phase (Part A) of the study through the participant decision visit or were unblinded or discontinued the study. Of these, 20 who went on to receive a single open-label booster dose of 50 ug mRNA-1273 (Part B) were selected for this preliminary analysis, with selection based on completion of their D15 visit assessments and immunogenicity sample availability. In the previously reported COVE phase 3 trial of mRNA-1273, 14711 (96.9%) participants completed both vaccinations with 100 µg mRNA-1273 in the blinded phase (Part A) of the study (11) and 60 participants at a single site were selected by site based on inclusion/exclusion criteria for Part C, to receive single open-label booster doses of 50 ug of mRNA-1273.351 (Part C, cohort 1) or mRNA-1273.211 (Part C, cohort 2) or 20 ug of mRNA-1273.351 (Part C, cohort 3). Only results for 50 µg booster doses of mRNA-1273 (selected subjects in Part B) or mRNA-1273.351 (Part C cohort 1) are available and reported here.

### Demographics and Characteristics

The baseline demographics of the group of participants who received the booster dose of the mRNA-1273.351 vaccine were generally similar to that for the group who received the booster dose of the mRNA-1273 vaccine (Table 1). Approximately half of the participants in both groups were male, and most were White and not Hispanic or Latino. The mean (range) age of the participants who received the booster dose of mRNA-1273.351 was 53.9 (27-70) years and for those who received mRNA-1273, 63.8 (38-76) years. The mean (SD) [range]) time between the second dose of mRNA-1273 and the booster dose of mRNA-1273 or mRNA-1273.351 was 201.0 (15.1) [177-226] and 187.1 (9.4 [168-198]) days, respectively.

**Table 1:**
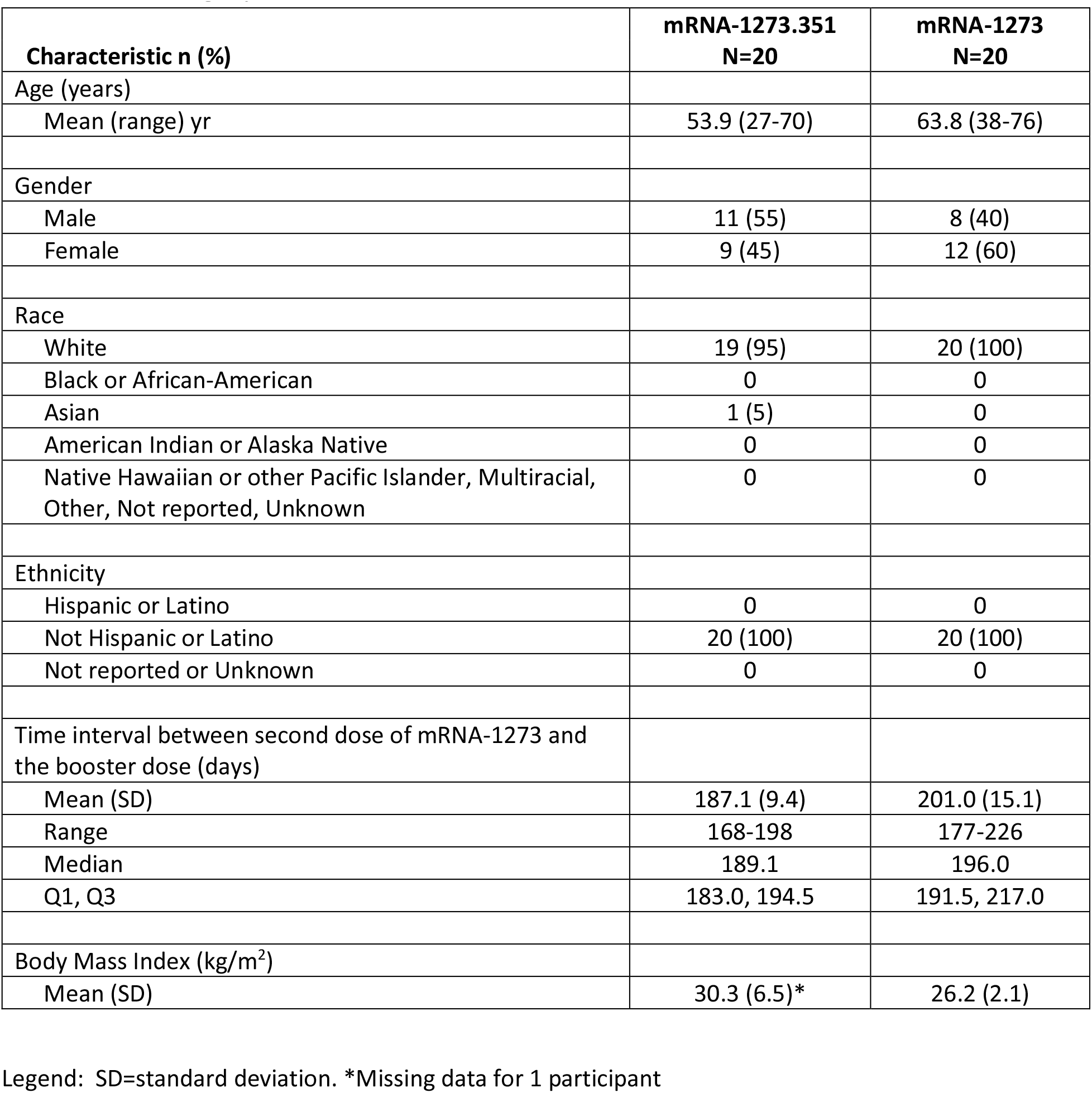
Demographics and characteristics

### Safety

With respect to safety, the percentages of participants with solicited local and systemic adverse events were similar in the group who received mRNA-1273.351 as a booster dose compared to those who received mRNA-1273 vaccine as a booster dose (Figure 2). The majority of solicited local and systemic adverse events were mild (grade 1) or moderate (grade 2). The frequency of any grade 3 solicited local or systemic adverse event was 15% (3 of 20 participants) after the booster dose of mRNA-1273 and 10.5% (2 of 19 participants) after the booster dose of mRNA-1273.351. There were no grade 4 solicited local or systemic adverse events. The most common solicited local adverse event was injection site pain after injection in both groups (68.4% for the mRNA-1273.351 vaccine and 90.0% for the mRNA-1273 vaccine) (9, 11). The most common solicited systemic adverse events after the booster dose of the mRNA-1273.351 vaccine were fatigue (36.8%), headache (36.8%), myalgia (31.6%) and arthralgia (21.1%). The most common solicited systemic adverse events after the booster dose of the mRNA-1273 vaccine were fatigue (70.0%), headache (55.0%), arthralgia (50.0%) and myalgia (45.0%). Fever was reported after the booster dose of mRNA-1273 in 3 of 20 participants (15%) but not after the booster dose of mRNA-1273.351 (0 of 19 participants). There were no serious adverse events reported in this study.

**Figure 2:**
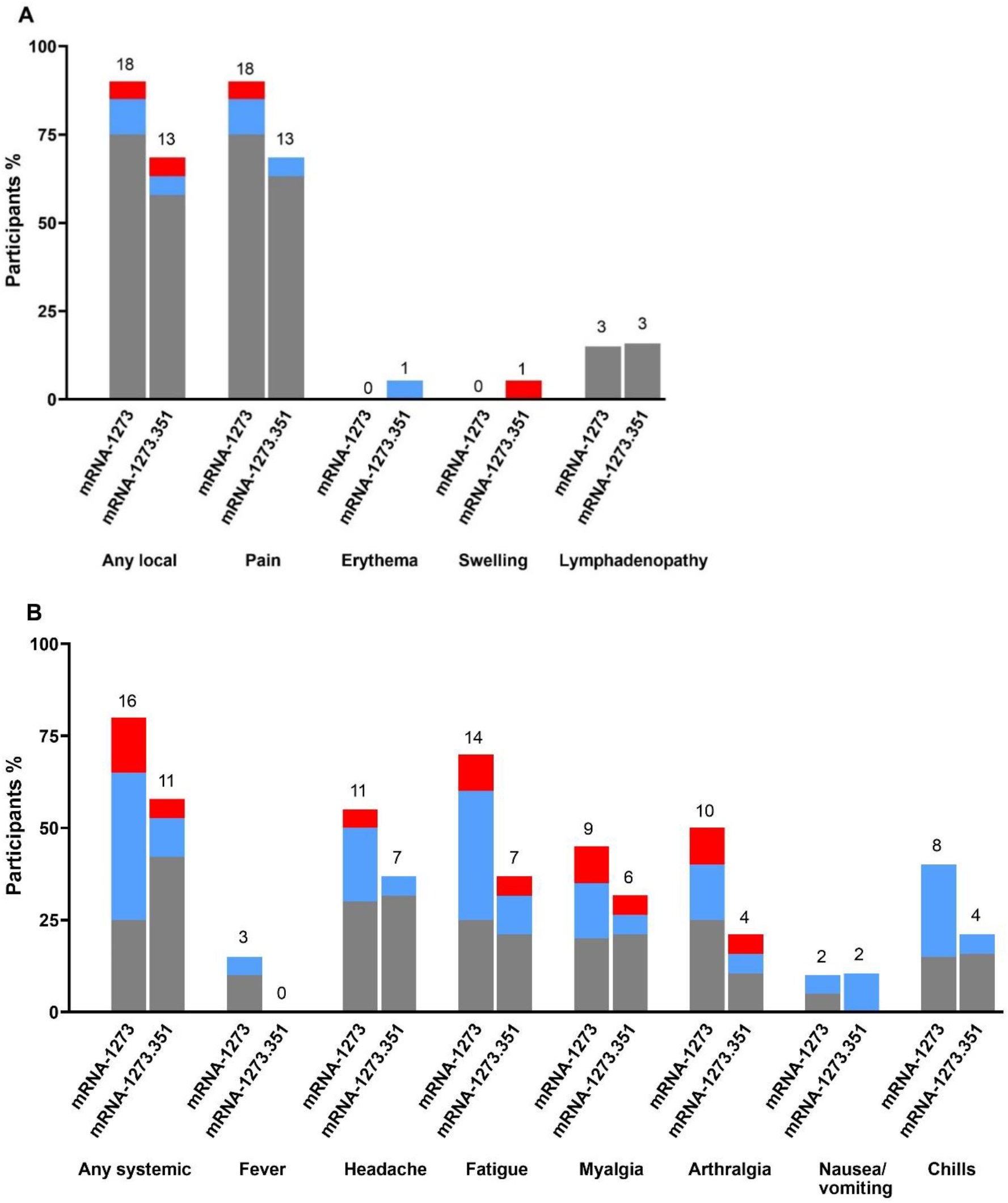
Solicited Local and Systemic Adverse Events Within 7 Days After Vaccination with a Booster Dose of mRNA-1273 or mRNA-1273.351. The percentage of participants who reported Grade 1 (gray), Grade 2 (blue), or Grade 3 (red) local **(A)** and systemic **(B)** adverse events is shown in the figure for the 20 participants who received a booster dose of mRNA-1273 and the 19 participants who received a booster dose of mRNA-1273.351. One participant who received mRNA-1273.351 did not report any results for solicited adverse reactions and was excluded from the analysis. The number above each bar shows the number of participants who reported the particular adverse event.

### Serum Neutralizing Antibody Titers Against Wild-type and Variant Viruses Prior to Booster Vaccinations

The preclinical SARS-CoV-2 PsVN assay was used to evaluate trial participant sera collected on the same day of vaccination with a booster dose of mRNA-1273 or mRNA-1273.351. While sera collected from these trial participants immediately after the primary series vaccinations have not yet been assessed, previous evaluations of mRNA-1273 vaccinated individuals at similar timepoints post-vaccination demonstrate that peak titers are ∼10^3^ ID_50_ (1-week post 2^nd^ dose titers). After a mean of 201.0 (15.1) [177-226] and 187.1 (9.4 [168-198]) days between the second dose of mRNA-1273 and the booster dose of mRNA-1273 or mRNA-1273.351 respectively, neutralizing antibody GMT against the original Wuhan-Hu-1 strain with the D614G mutation (referred to as wild-type, D614G throughout the text and figures) in these individuals have decreased to 198 in the Part B cohort and 304 in the Part C cohort 1 (Figure 3 A/C, D1 Pre 3^rd^ dose). However, neutralization of the B.1.351 and P.1 variants was further reduced. GMT levels of neutralizing antibodies of 27 or 40, and 30 or 47 were measured against B.1.351 or P.1 in the Part B and Part C cohort 1 samples respectively. In addition, approximately 50% of samples from both cohorts fell below the assay LLOQ against B.1.351 and P.1 viral variants.

**Figure 3A and B:**
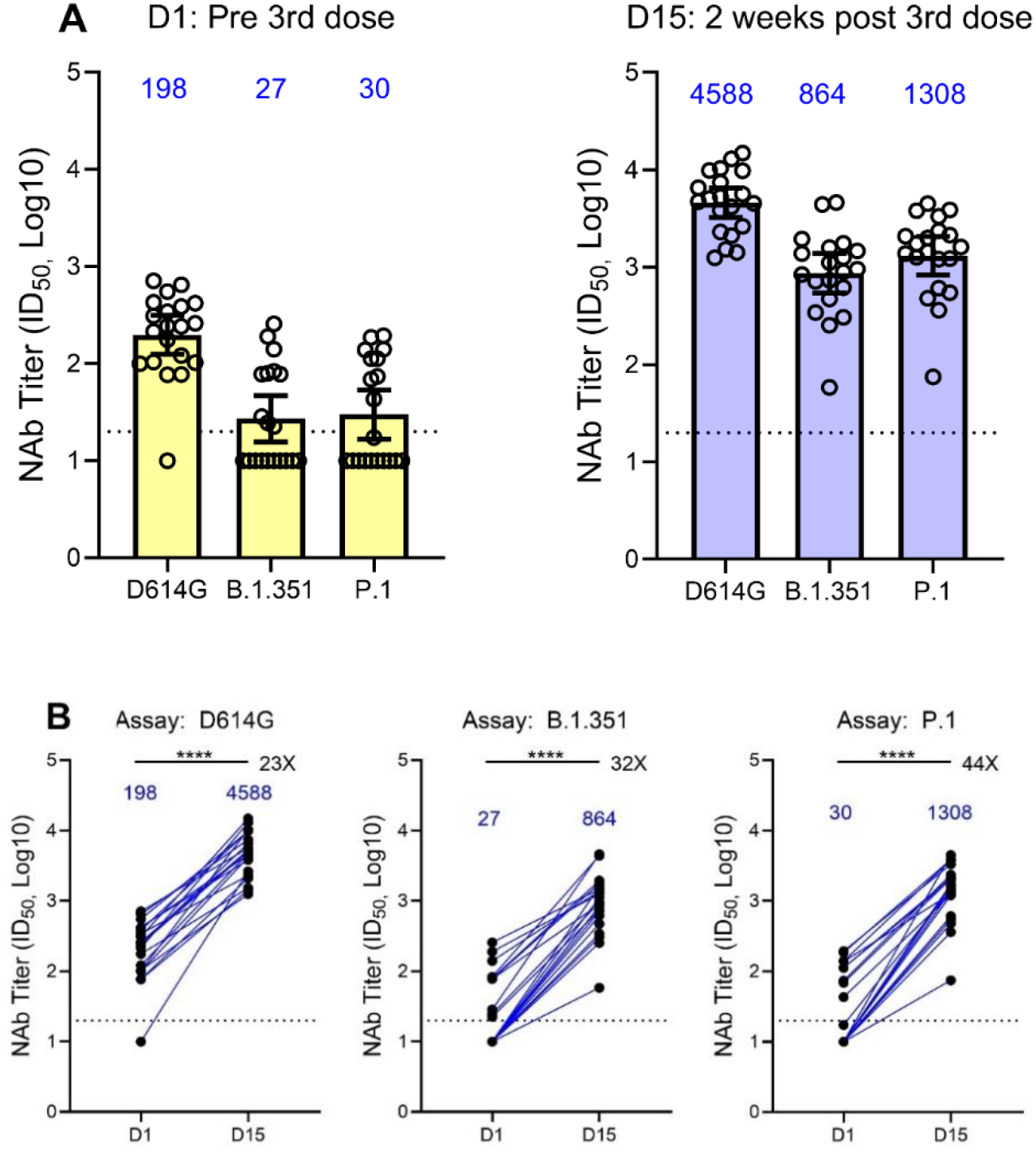
Immunogenicity After Boosting with Booster Dose of 50 µg of mRNA-1273.

**Figure 3C and D:**
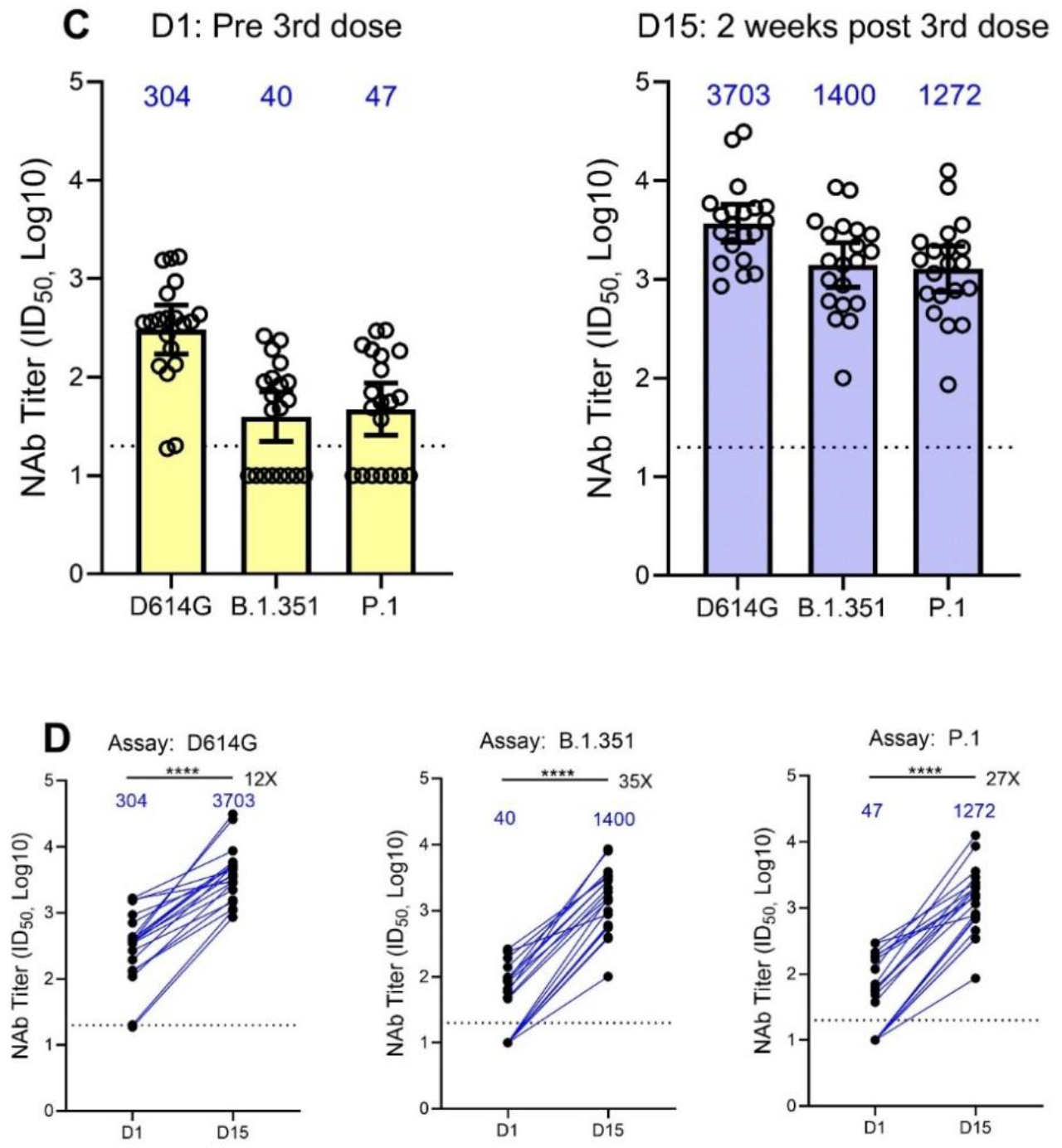
Immunogenicity After Boosting with 50 µg of mRNA-1273.351. Neutralization of recombinant SARS-CoV-2 VSV-based pseudoviruses (D614G, B.1.351 and P.1) by serum from participants before (D1) and 15 days after boosting (D15) with 50 µg of mRNA-1273 **(A and B)** and mRNA-1273.351 **(C and D)**. The geometric mean neutralizing antibody titer is denoted by the top of the box and the 95% confidence intervals are shown by the brackets. The titers for individual participants are shown by the circles. The fold increases for day 15/day 1 are shown above the bars. The horizonal dotted lines indicate the lower limit of quantification (LLOQ). **** = p<0.0001 by the Wilcoxon matched-pairs signed rank test.

### Neutralization after vaccination with a booster dose of mRNA-1273 or mRNA-1273.351 in individuals previously vaccinated with mRNA-1273

To evaluate the ability of mRNA-1273 or mRNA-1273.351 to boost pre-existing immunity and increase neutralization against both the wild-type, B.1.351, and P.1 virus, P201 trial participants were immunized with 50 µg mRNA-1273 (Figure 1, Part B), while participants from P301 were enrolled into P201 Part C and dosed with 50ug mRNA-1273.351 (Figure 1, Part C cohort 1). Sera collection occurred 2 weeks after vaccination with each booster vaccine, and neutralization assessments were performed. Additional arms of this trial are ongoing to evaluate boosting with a multivalent mRNA-1273.211 that contains both mRNA-1273 and mRNA-1273.351 in a 1:1 mix.

Neutralization titers were measured in the D614G, B.1.351, and P.1 preclinical SARS-CoV-2 PsVN assays prior to the boost (D1) and two weeks after (D15). Boosting with mRNA-1273.351 or mRNA-1273 increased neutralization antibody titers (Figure 3). Boosting with mRNA-1273 increased PsVN GMTs to 4508, 864, and 1308 in the respective D614G, B.1.351, and P.1 assays (Figure 3A), representing a 23, 32, and 44-fold rise in neutralization against each respective virus (Figure 3B). Boosting with mRNA-1273.351 increased PsVN neutralizing titers to 3703, 1400, and 1272 (Figure 3C), and represented a 12, 35, and 27-fold increase in neutralization of each respective virus (Figure 3D). Importantly, vaccination with a booster dose of mRNA-1273 or mRNA-1273.351 resulted in the detection of robust neutralization titers against both wild-type and variant viruses after the boost in all participants. In addition, titers against B.1.351 and P.1 assay now approach or exceed 10^3^ ID_50_ GMT, the previous peak titers in the wild-type assay (Figure 4E).

**Figure 4:**
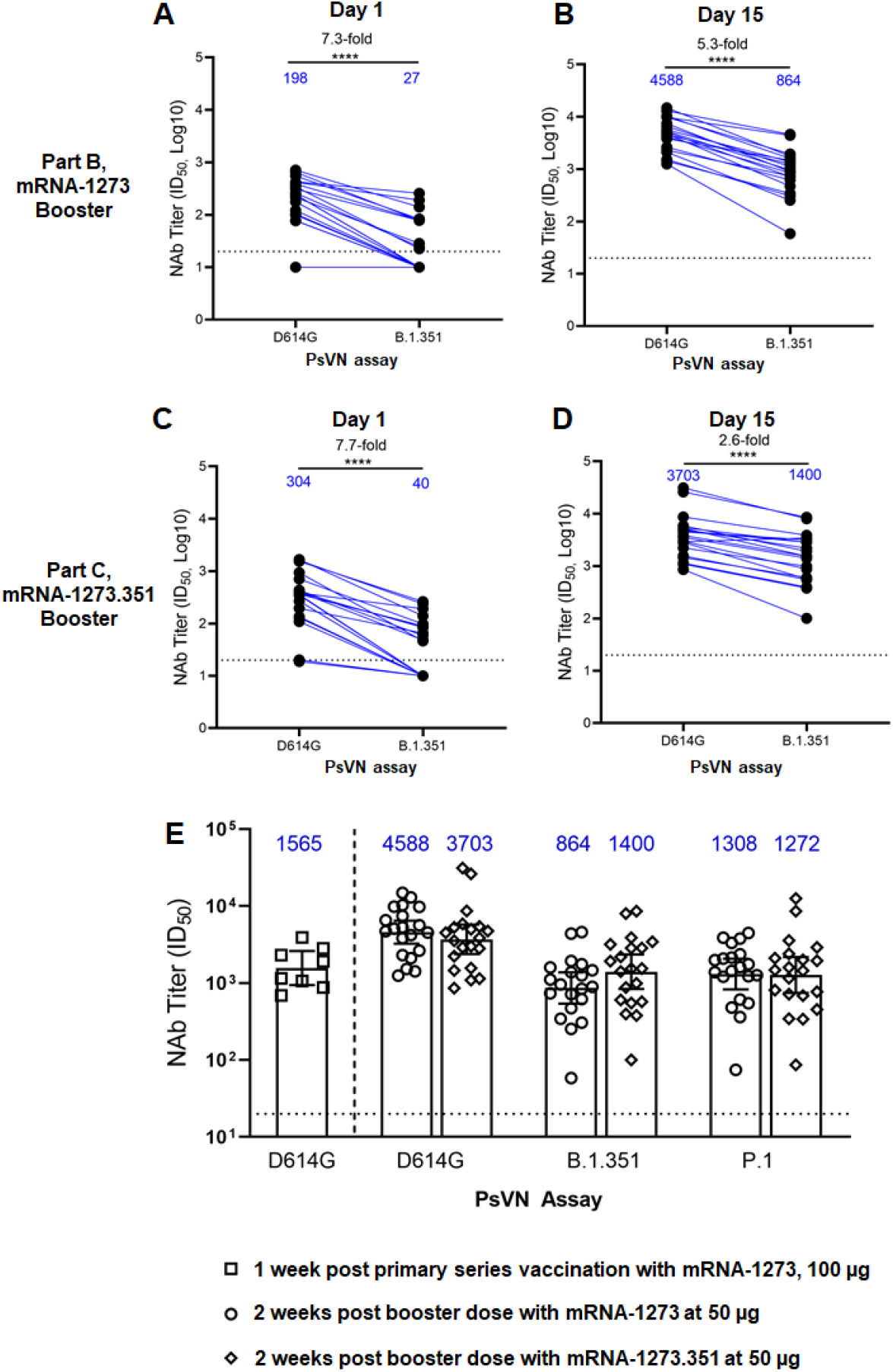
Neutralization of D614G and B.1.351 SARS-CoV-2 pseudoviruses by serum from mRNA-1273 or mRNA-1273.351 boosted trial participants on D1 (pre-boost) and D15 (post-boost) Sera was collected from trial participants before and after vaccination with a 50 µg booster dose. mRNA-1273 boosted subjects before (**A**) and after (**B**) the boost. mRNA-1273.351 boosted subjects before (**C**) and after (**D**) the boost. Reference titers in the D614G assay for mRNA-1273 vaccinated subjects after the primary vaccination series of mRNA-1273, versus titers measured in the PsVN assays two weeks after boost with mRNA-1273 or mRNA-1273.351 (**E**). Fold-difference in neutralization between the D614G and B.1.351 VSV PsVN assays are indicated. **** = p<.0001 by the Wilcoxon matched-pairs signed rank test. Blue text indicates the geometric mean titers, which is listed as text above each plot. The horizontal dotted lines indicate the lower limit of quantitation for Nab titer at 10 ID_50_. Results from individual participants are represented as dots on each figure, with lines connecting the D614G and B.1.351 neutralization titers.

### A mRNA-1273.351 booster more effectively narrows the gap in neutralizing titers between the wild-type and B.1.351 viruses, versus mRNA-1273

Prior to vaccination with the booster dose, neutralization titers were 7.3 and 7.7-fold reduced versus the B.1.351 virus in the Part B and Part C cohort 1 participants respectively (Figure 4 A, B). Two weeks after the booster dose, a 5.3 and 2.6-fold difference between wild-type and B.1.351 neutralization was measured, and importantly all samples were able to neutralize all viruses (Figure 4 C, D). Importantly, the variant mRNA-1273.351 booster was able to increase titers against the B.1.351 virus to higher levels versus a boost with mRNA-1273 (1400 versus 864 ID_50_ GMT), and was able to further close the gap of neutralization between the two assays to 2.6-fold. Equally important, titers against the variant viruses approach peak titers measured after the primary series vaccination with mRNA-1273, from an analysis of phase 1 trial samples measured in the homologous wild-type assay (Figure 4E) (8).

## Discussion

This is a preliminary evaluation of mRNA-1273 and mRNA-1273.351 given as boosters to individuals that had been vaccinated 6.2 to 6.7 months previously with mRNA-1273 in an amended phase 2 clinical trial of mRNA-1273. The safety profiles following single injections of 50 µg of mRNA-1273 (Part B) or mRNA-1273.351 (Part C cohort 1) were evaluated, and although similar to those observed after a second dose of mRNA-1273 in the previously reported phase 2 and phase 3 studies, some differences were seen between the two booster vaccines. Vaccination with both mRNA-1273 and mRNA-1273.351 boosters elicited higher neutralizing titers against the wild-type original strain and comparable titers against the B.1.351 and P.1 variants versus peak titers observed after the primary series vaccinations as measured against the wild-type virus (Figure 4E), suggesting that immune memory was induced by mRNA-1273 priming. Additionally, the mRNA-1273.351 booster appeared to be more effective at increasing neutralization against the B.1.351 variant than a boost with mRNA-1273. The study is ongoing and evaluation of boosting of clinical trial participants with the multivalent mRNA-1273.211 is underway.

In this preliminary report, boosters of 50 µg of mRNA-1273 or mRNA-1273.351 given to individuals who were previously vaccinated with two doses of mRNA-1273 elicited acceptable safety profiles. Solicited local and systemic adverse events after a booster dose of mRNA-1273.351 or mRNA-1273 were similar. The majority of events were mild (grade 1) or moderate (grade 2) in severity, and grade 3 events occurred with a frequency of 15% in Part B and 10% in Part C. No grade 4 events were reported. For both boosters, the most commonly reported solicited local adverse event was injection site pain and reported systemic events included fatigue, headache, myalgia and arthralgia, consistent with safety profiles seen in the phase 2 and 3 studies (9, 11). It should be noted that limited adverse events of fever post-vaccination were reported in the Part B (mRNA-1273; 15%) but not the Part C (mRNA-1273.351; 0%) participants, similar to those reported after the second mRNA-1273 vaccinations in the phase 2 and phase 3 clinical trials. Importantly, in this small study, the booster dose of mRNA-1273.351 appears to be tolerated (solicited local and systemic adverse events) at least as well if not better than a booster dose of mRNA-1273.

Sera from these participants collected prior to the boost (D1) and two weeks after (D15) were evaluated in a preclinical SARS-CoV-2 PsVN neutralization assay against wild-type virus.

Prior to the boost, neutralization titers of 198 and 304 in the Part B and Part C cohorts remained significant, at levels predicted to be protective against the original Wuhan-Hu-1 isolate (19). These results are consistent with those reported in a lentiviral PsVN assay, where monitoring of sera neutralization titers was performed up to 6 months after the second dose of mRNA-1273 (16). In that evaluation, the lentiviral PsVN titer was 57 ID_50_ GMT in participants who were 56 to 70 years old 180 days after the second dose of mRNA-1273.

In our evaluation of Parts B and C participant sera, virus neutralization assays were performed against B.1.351 and P.1 variants. Both are VOCs and contain the key receptor binding domain (RBD) mutations K417T/N, E484K, N501Y present in the mRNA-1273.351 variant vaccine. Significantly reduced levels of neutralization were measured against both the B.1.351 and P.1 variant viruses in samples collected prior to the booster vaccination, with ∼50% of titers falling below the assay lower limit of quantification. Although a correlate of protection has not yet been established for SARS-CoV-2 infection or COVID-19 disease, lack of detectable neutralization against these variants may be indicative of waning immunity, especially against VOCs.

Boosting these trial participants with mRNA-1273 and mRNA-1273.351 both substantially increased neutralization titers against the wild-type, and B.1.351 and P.1 variant viruses (Figure 3). Neutralizing titers against the wild-type virus exceeded the peak titers measured after the primary series in separate studies (15, 16), indicating the induction of immune memory, and titers against the B.1.351 and P.1 viruses increased to a level very similar to those previously measured against the wild-type assay where ∼10^3^ ID_50_ GMT values were measured (15). Furthermore, all trial participants, including those that had undetectable titers against the variant viruses, had robust neutralization titers in both the wild-type and variant assays two weeks after the booster. A boost with mRNA-1273.351 appeared to be more effective at neutralization of the B.1.351 virus than a boost with mRNA-1273, evidenced by the higher mean GMT levels in the Part C cohort 1 participants (1400) than the GMT Part B participants (864) against the B.1.351 virus. Additionally, the difference between the wild-type and B.1.351 assays at day 1 dropped from 7.7-fold prior to the boost with mRNA-1273.351 to 2.6-fold at 15 days after the boost. Further reductions in the differences between the two assays may be found in samples collected from these participants at later timepoints, as the kinetics of the neutralizing antibody responses to the new epitopes in S-2P.351 protein encoded by mRNA-1273.351 may be different from the epitopes shared between the two immunogens. Strong homologous responses, both in terms of absolute geometric mean titer and geometric mean fold rise, assessed by the same strain in the assay used in the vaccine, were seen regardless of vaccine strain. Response to the wild-type virus was highest with a boost of mRNA-1273 and response to B.1.351 was highest with mRNA-1273.351. In addition, heterologous responses, against variants when prototype vaccine was used to boost or against prototype after the mRNA-1273.351 booster were also seen. This supports the development of a variant vaccine as a booster dose to prevent infection caused by variant strains.

This phase 2 study is ongoing, and data from additional arms, including an evaluation of a 50 ug booster dose of the multivalent mRNA-1273.211 vaccine comprised of a 1:1 mix of mRNA-1273 and mRNA-1273.351 in Part C cohort 2, will be reported later. This additional arm was designed to evaluate the effectiveness of the multivalent vaccine as a booster and its effectiveness at broadening the immune response to provide better neutralization against both wild-type virus and variants. In addition, sera collected on day 57 after the primary vaccination series from all these trial participants will be analyzed in the preclinical SARS-CoV-2 PsVN assay to allow for direct comparison of peak titers prior to the boost versus neutralizing titers elicited from each booster dose, allowing for better evaluation of the level of waning immunity seen in these participants and further demonstration of the benefit from the booster dose. Global surveillance for the emergence of additional SARS-CoV-2 VOCs and efforts to test the neutralization of VOCs by mRNA-1273 vaccinee sera as well as sera from these boosted trial participants is also ongoing. If additional variants emerge that reduce the neutralization capacity of 2-dose mRNA-1273 or 2-dose mRNA-1273 + booster regimen, alternative mRNA vaccine designs may be developed and evaluated clinically. As variants are likely to emerge and continue to co-circulate globally, it is expected that a multivalent mRNA vaccine design will likely be most effective at increasing cross-variant protection and will also afford the opportunity to mix and match which variant designs are included in the vaccine to respond to the continued evolution of the SARS-CoV-2 virus and address potentially changing global needs.

There are some limitations related to the preliminary analysis of this study. First, the results presented here are based on a limited sample size. The trial is ongoing, and key evaluations of the multivalent mRNA-1273.211 arm of the trial have not yet been performed. The neutralization assay used in the evaluations of these pre-boost and two-week post-boost samples is a preclinical SARS-CoV-2 PsVN assay, and although this assay has consistently been used to evaluate the impact on neutralization against variant viruses, the assay has not been qualified (16). All samples from this trial will be evaluated in qualified assays and the results will be reported at a later date. As analysis of sera for the trial participants collected on day 57 after the primary series of mRNA-1273 has not yet been performed in this preclinical assay, it cannot yet be definitively determined how the neutralization titers after the 3^rd^ dose will compare to peak measurements after the primary vaccination series. Further, evaluations of the sera collected two weeks after the booster have not yet been performed against other VOC, including B.1.526, B.1.427/B.1.429, or B.1.617; therefore, it remains to be seen if the breadth of protection against other variants has increased after the booster was given. Also, as a correlate of protection for neutralizing antibodies has not yet been established for SARS-CoV-2, it can’t be definitively determined from these preliminary results whether the significant neutralization titers elicited from both booster vaccines would be protective against B.1.351 or P.1, although it seems probable. Finally, because the participants in this study were originally enrolled from two different clinical trials (Part B; phase 2 mRNA-1273 study and Part C, Cohort 1; phase 3 mRNA-1273 COVE study), a comparison of the results of a booster dose of mRNA-1273.351 with those of mRNA-1273 should be interpreted with caution.

The emergence of SARS-CoV-2 variants and the ability of the virus to partially overcome natural or vaccine-induced immunity has served as a call to action. Not only are continued vaccination efforts needed to prevent the emergence of future VOCs, but also strategies for SARS-CoV-2 vaccine research and development are needed that can enhance the level of protection against key VOCs, should they arise. The mRNA platform approach against SARS-CoV-2 VOCs in this trial appears to be effective at boosting antibody levels when applied as a booster dose, with mitigation of the reductions in neutralization seen against the B.1.351 and P.1 lineages. The mRNA platform allows for rapid design of vaccine antigens that incorporate key mutations, allowing for faster development of future alternative variant-matched vaccines should they be needed. The vaccine designs evaluated in this clinical study demonstrate the ability to boost immunity to titers that likely exceed those that peak after the primary vaccination series against both the wild-type virus and variants, and also demonstrate the potential of the mRNA-1273.351 booster to close the gap between neutralization of the wild-type virus and the B.1.351 variant. In the future, additional VOC designs can be rapidly developed, evaluated, and deployed if needed to address the evolving SARS-CoV-2 virus.

## Data Availability

Data sharing statement: Moderna is committed to sharing data supporting the findings of eligible studies. The results of this study are preliminary and the study is ongoing. Access to patient-level data and supporting clinical documents with qualified external researchers may be available upon request once the trial is complete.

## Acknowledgements

We thank Frank J Dutko and Joanne E Tomassini, Moderna consultants, for contributions to writing the manuscript, and Michael Brunner and Dr. Michael Whitt for kind support on recombinant VSV-based SARS-CoV-2 pseudovirus production.

